# Zero-Shot Prompting is the Most Accurate and Scalable Strategy for Abstracting the Mayo Endoscopic Subscore from Colonoscopy Reports Using GPT-4

**DOI:** 10.1101/2024.03.22.24304745

**Authors:** Richard P. Yim, Vivek A. Rudrapatna

**Author notes:** **<u>Correspondence:</u>** Vivek A. Rudrapatna, MD, PhD, 490 Illinois St, Ste 21E, San Francisco, CA 94143. **<u>Writing Assistance:</u>** None. **<u>Author Contributions:</u>** *Yim:* Study concept and design; data extraction; analysis and interpretation of data; drafting of manuscript; critical revision of the manuscript for important intellectual content *Rudrapatna:* Study concept and design; data extraction; analysis and interpretation of data; drafting of manuscript; study supervision; critical revision of the manuscript for important intellectual content.

## Abstract

**Introduction:** Large-language models can help extract information from clinical notes, making them potentially useful for research in ulcerative colitis. However, it remains unclear if these models will scale well in practice.

**Methods:** We analyzed the performance and cost of programmatically using GPT-4 to abstract Mayo endoscopic subscores (MES) from 499 colonoscopy reports using different prompting strategies.

**Results:** Zero-shot prompting, where GPT-4 is instructed without examples, was most accurate (83.55%) *and* cost-effective ($0.097/note).

**Discussion:** Using GPT-4 to automatically curate the MES and other variables is a practical strategy for quantifying UC activity and measuring improvements to clinical care.

## Introduction

The Mayo endoscopic subscore (MES) is a core measure of ulcerative colitis (UC) activity (1), but is not always explicitly documented in colonoscopy reports. Thus, clinical studies that use the MES frequently require manual review of these reports to abstract these scores from free-text descriptions. Large language models like GPT-4 have shown promise in their ability to extract information from clinical notes Prior studies have used the more user friendly, chatbot interface to interact with these models. However, these models can also be used in a programmatic fashion, raising the possibility of being natively deployed within electronic health record (EHR) systems to dynamically maintain disease registries, optimize study recruitment, and support quality improvement.

As a next step, we studied the scalability and cost-effectiveness of using GPT-4 to automate the extraction of the modified MES. We hypothesized that more sophisticated, *n-shot* and iterative-style prompts would yield more accurate results despite higher costs associated with this strategy.

## Methods

We utilized an existing set of 499 annotated colonoscopy reports sourced from two hospitals in California 217 were from San Francisco General Hospital (SFGH), a safety net hospital, and 282 from the University of California, San Francisco Health, a tertiary care hospital. These reports were annotated based on 1) their suitability for MES scoring (e.g. clear diagnosis of UC, surgically unaltered anatomy), and, 2) the modified MES (4) if appropriate.

We developed two generic conversation templates for zero-shot and *n*-shot prompting to programmatically interact with GPT-4-turbo via LangChain (5), a framework that enables context-rich prompts, and UCSF Versa, a PHI-compliant programmatic interface with GPT-4. (See **Table 1, Supplemental Digital Content 1**, for precise prompt templates and protocol texts.) We refer to *n*-shot prompting as providing *n* colonoscopy reports per Mayo score plus a non-Mayo scorable report in addition to the scoring protocol; zero shot refers to providing only the scoring protocol. For these templates we also studied the performance of GPT-4 with prompts that asked it to not only include the MES, but an *explanation* as well. We also provided GPT-4 with a *parsed* variation of colonoscopy report for UCSF and SFGH centers where only the main relevant text of the colonoscopy procedure report was provided as opposed to the colonoscopy report text in its entirety, which includes extraneous text strings. See **Method Details, Supplemental Digital Content 1**, for additional explanation.

**Table 1.** Performance results, classification error, and cost are shown across each prompt template variation and data variation. Bolded text means higher is better; italicized text means lower is better. Best numeric values along each measurement have been underlined. Table has been partitioned according to template variation.

| Prompt Template Variation | Zero-shot | Zero-shot | Zero-shot | Zero-shot | Zero-shot | Zero-shot | Zero-shot | Zero-shot | Zero-shot, two-task | Zero-shot, two-task | One-shot | One-shot | Four-shot | Four-shot |
| --- | --- | --- | --- | --- | --- | --- | --- | --- | --- | --- | --- | --- | --- | --- |
| Center | UCSF | UCSF | SFGH | SFGH | UCSF | UCSF | SFGH | SFGH | UCSF | SFGH | UCSF | SFGH | UCSF | SFGH |
| With Parsing | Trimmed | Trimmed | Trimmed | Trimmed | Original | Original | Original | Original | Trimmed | Trimmed | Trimmed | Trimmed | Trimmed | Trimmed |
| With Explanation | Yes | No | Yes | No | Yes | No | Yes | No | Yes | Yes | Yes | Yes | Yes | Yes |
| Accuracy* | **<u>83.55%</u>** | **82.16%** | **77.17%** | **78.11%** | **83.11%** | **81.45%** | **73.15%** | **75.86%** | **<u>83.95%</u>** | **78.59%** | **<u>79.23%</u>** | **73.04%** | **<u>75.93%</u>** | **73.82%** |
| Precision* | **<u>87.33%</u>** | **86.94%** | **81.83%** | **82.72%** | **87.01%** | **85.37%** | **79.29%** | **81.37%** | **<u>87.17%</u>** | **82.57%** | **<u>82.78%</u>** | **80.51%** | **<u>80.46%</u>** | **77.88%** |
| Recall* | **<u>85.11%</u>** | **84.75%** | **79.26%** | **80.18%** | **83.69%** | **82.27%** | **75.58%** | **78.34%** | **<u>85.11%</u>** | **81.11%** | **<u>75.09%</u>** | **74.06%** | **<u>70.61%</u>** | **74.62%** |
| F1 score* | **<u>85.62%</u>** | **85.37%** | **79.51%** | **80.67%** | **84.51%** | **83.12%** | **76.42%** | **78.81%** | **<u>85.42%</u>** | **81.16%** | **<u>76.42%</u>** | **75.88%** | **<u>71.95%</u>** | **75.64%** |
| Scorable Accuracy | **92.95%** | **<u>93.66%</u>** | **92.24%** | **92.57%** | **91.81%** | **90.28%** | **92.47%** | **91.81%** | **<u>92.34%</u>** | **92.33%** | **82.35%** | **<u>84.28%</u>** | **78.76%** | **83.33%** |
| Severity Agreement<sup>†</sup> | **90.73%** | **<u>91.39%</u>** | **82.12%** | **82.78%** | **89.40%** | **90.73%** | **78.81%** | **82.12%** | **<u>90.73%</u>** | **82.78%** | **92.52%** | **80.95%** | **<u>93.33%</u>** | **82.22%** |
| Under Classification(1) | **<u>7.28%</u>** | **8.61%** | **10.60%** | **11.92%** | **8.61%** | **9.27%** | **14.57%** | **12.58%** | **<u>3.97%</u>** | **7.95%** | **<u>9.52%</u>** | **15.66%** | **<u>10.37%</u>** | **12.59%** |
| Under Classification(2) | **<u>0.66%</u>** | **<u>0.66%</u>** | **1.32%** | **1.32%** | **<u>0.66%</u>** | **<u>0.66%</u>** | **1.32%** | **1.32%** | **<u>0.66%</u>** | **<u>0.66%</u>** | **<u>0.68%</u>** | **1.36%** | **<u>0.74%</u>** | **<u>0.74%</u>** |
| Average cost per note | **\$0.097** | **<u>\$0.096</u>** | **\$0.129** | **\$0.128** | **\$0.108** | **\$0.107** | **\$0.152** | **\$0.151** | **<u>0.148</u>** | **\$0.267** | **\$0.499** | **<u>\$0.280</u>** | **<u>\$0.803</u>** | **\$1.485** |

\*Computed using weighted-average of metric for each class.
<sup>†</sup> Severe UC as measured by having an MES of 2 or 3.

## Results

Zero-shot prompts on trimmed notes produced the best performing results consistently on both UCSF (81.45-83.55% weighted average accuracy) and SFGH (73.15-78.11% weighted average accuracy) reports (**Table 1**). We found that that *n*-shot prompting actually decreased classification performance across multiple metrics, rejecting our hypothesis that providing examples would improve GPT-4’s performance. Further, the cost of *n*-shot prompting is prohibitively more expensive on average (e.g., more than *8 times* the cost for UCSF reports and *11 times* for SFGH reports between *n*-shot and zero-shot prompting *per note*). For Mayo scorable accuracy, whether an MES can be assigned to the colonoscopy report, we find that GPT-4 performs very well for zero-shot prompt templates (accuracy 90.28-93.66%), where *n*-shot prompting reduced its accuracy (best score at 84.28%). We also studied results for splitting Mayo scorable reports and MES separately (“zero-shot, two-task prompting”) but performance gains were negligible (**Table 1**).

With respect to prompt variations such as parsing colonoscopy reports and soliciting explanations for MES values, across all strata we find that the greatest difference in performance is 4.02% for trimming and -3.31% for requiring an explanation (**Table 2**). In particular, parsing the text generally increases performance across all measures and decreases under classification as well. Interestingly, we find that across standard statistical learning metrics, the UCSF data shows an increase in performance when requiring explanation of the MES and a decrease in performance for SFGH data although the magnitude of these differences is minimal (worst difference in magnitude amongst accuracy, precision, recall and F1-score is 2.76%).

**Table 2.**
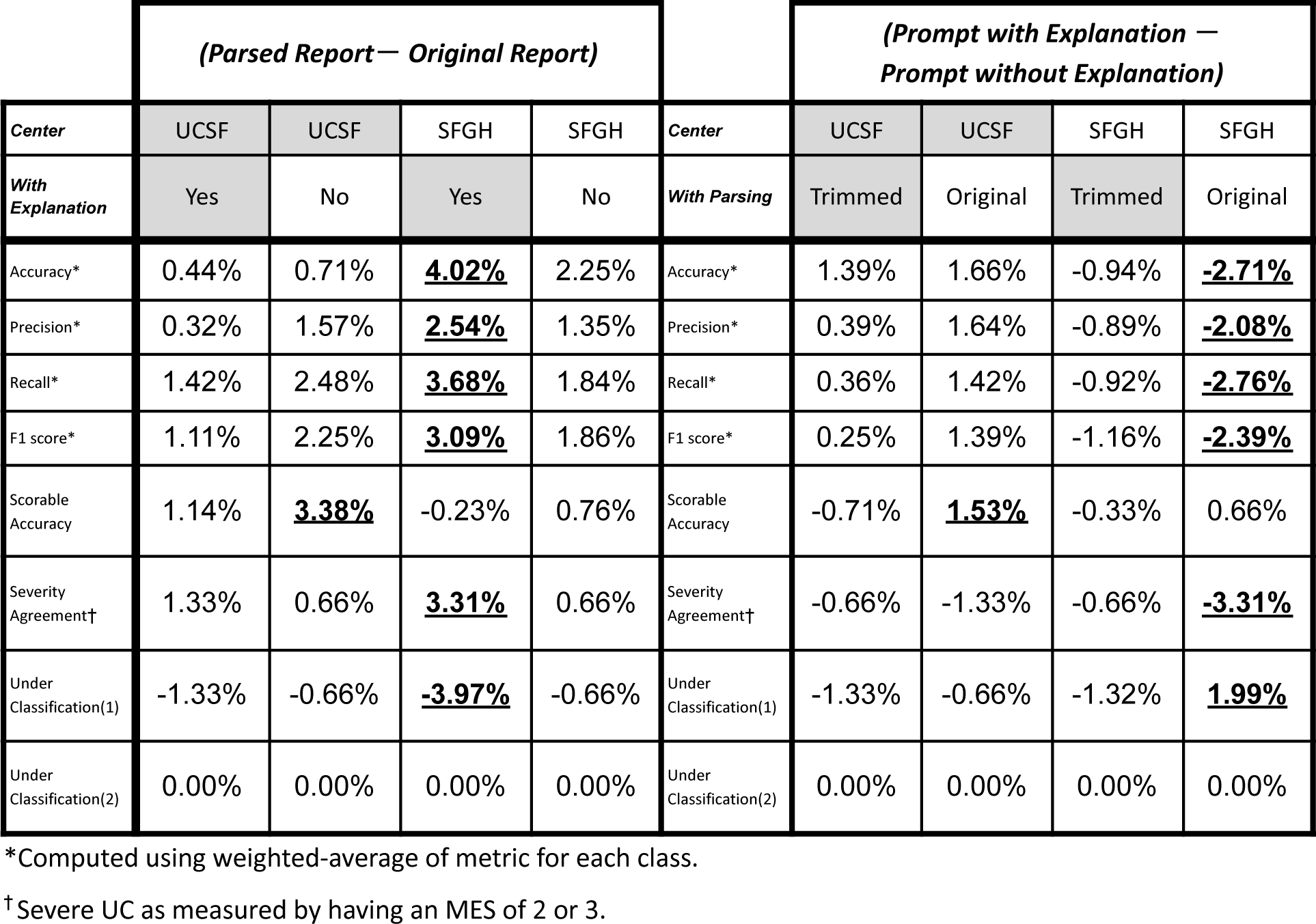
DIfferences in performance stratified on parsing colonoscopy report text (*Parsed Report - Original Report*) and requiring explanation (Prompt with *Explanation - Prompt without explanation*).

| *(Parsed Report — Original Report)* |  |  |  |  | *(Prompt with Explanation — Prompt without Explanation)* |  |  |  |  |
| --- | --- | --- | --- | --- | --- | --- | --- | --- | --- |
| *Center* | UCSF | UCSF | SFGH | SFGH | *Center* | UCSF | UCSF | SFGH | SFGH |
| *With Explanation* | Yes | No | Yes | No | *With Parsing* | Trimmed | Original | Trimmed | Original |
| Accuracy* | 0.44% | 0.71% | **<u>4.02%</u>** | 2.25% | Accuracy* | 1.39% | 1.66% | -0.94% | **<u>-2.71%</u>** |
| Precision* | 0.32% | 1.57% | **<u>2.54%</u>** | 1.35% | Precision* | 0.39% | 1.64% | -0.89% | **<u>-2.08%</u>** |
| Recall* | 1.42% | 2.48% | **<u>3.68%</u>** | 1.84% | Recall* | 0.36% | 1.42% | -0.92% | **<u>-2.76%</u>** |
| F1 score* | 1.11% | 2.25% | **<u>3.09%</u>** | 1.86% | F1 score* | 0.25% | 1.39% | -1.16% | **<u>-2.39%</u>** |
| Scorable Accuracy | 1.14% | **<u>3.38%</u>** | -0.23% | 0.76% | Scorable Accuracy | -0.71% | **<u>1.53%</u>** | -0.33% | 0.66% |
| Severity Agreement† | 1.33% | 0.66% | **<u>3.31%</u>** | 0.66% | Severity Agreement† | -0.66% | -1.33% | -0.66% | **<u>-3.31%</u>** |
| Under Classification(1) | -1.33% | -0.66% | **<u>-3.97%</u>** | -0.66% | Under Classification(1) | -1.33% | -0.66% | -1.32% | **<u>1.99%</u>** |
| Under Classification(2) | 0.00% | 0.00% | 0.00% | 0.00% | Under Classification(2) | 0.00% | 0.00% | 0.00% | 0.00% |

\*Computed using weighted-average of metric for each class.
† Severe UC as measured by having an MES of 2 or 3.

## Discussion

This is among the first few studies to use an LLM in a programmatic fashion to extract study variables from clinical notes. We found that the most accurate *and* scalable prompting strategy is conveniently the most simple when it comes to producing MES scores from colonoscopy reports. Zero-shot prompts are not only easy to implement, but cost effective. GPT-4 proves itself to be reasonably effective at being able to simultaneously determine whether a colonoscopy procedure report is Mayo scorable, and providing an MES when it is. Further, we find that *n-shot* prompting is unreliable both in performance and cost.

Beyond template variations of zero-shot, *n*-shot, and zero-shot, two-task prompting, our study explores prompt parameter interactions in GPT-4 performance that are currently absent in the literature (e.g., explanation requirement and text parsing). Further, our study explores the possibility of the generalizability of LLM information extraction across different centers. The primary limitation of our study then is a sophisticated endpoint. For instance, although the colonoscopy report distribution for IBD patients is representative across UCSF and SFGH centers, we have limited class representation for more severe MES graded UC (Mayo scores 2 and 3 in particular). Other studies in the literature explore continuously valued endpoints as well multidimensional endpoints extracted from clinical text (6,7).

However, these studies focus primarily on the performance of GPT-4 and LLMs on clinical notes. There has been little consideration and commentary on prompt engineering *and* consequently the costs of deployment—in other words, the practicality of deploying GPT-4 for other studies. Deploying four-shot prompts on 282 parsed UCSF colonoscopy reports, requiring GPT-4 to produce an explanation, comes out to an average total cost of $418.77. There are thousands of colonoscopy procedure reports for IBD patients at our medical centers, but billions of notes across all diseases and patients in the US (8). Generative AI is and will be very expensive to deploy across all clinical areas and target variables. While LLMs like GPT-4 will enable retrospective information extraction, and consequently new observational studies using EHR data, we strongly advise clinical researchers to be mindful of various prompting strategies and their costs.

## Data Availability

All data is PHI-compliant and is not to be publicly available.

## Data Acknowledgement

The authors thank UCSF Academic Research Services for technical support related to enabling software in a secure, PHI compliant environment; UCSF AI Tiger Team for facilitating and managing access to Versa API (UCSF secure access to Microsoft Azure, OpenAI Large language Models); and the Chancellor’s Task Force for Generative AI.

