## Supplemental Document for "Zero-Shot Prompting is the Most Accurate and Scalable Strategy for Abstracting the Mayo Endoscopic Subscore from Colonoscopy Reports Using GPT-4"

**Supplemental Digital Content 1**

**Method Details**

### *Cohort Details*

The SFGH colonoscopy report cohort consists of 30.4% Mayo scorable colonoscopy reports---not all colonoscopies are UC related; 18% have an MES of 0; 11.5% with an MES of 1, 30.9% with an MES of 2 and 9.2% with an MES of 3. The UCSF colonoscopy report cohort consists of 46.5% Mayo scorable notes; 13.8% with an MES of 0; 21.6% with an MES of 1; 9.2% with an MES of 2; and 8.9% with an MES of 3.

### *Data Parsing*

Colonoscopy reports from these two centers have idiosyncrasies with respect to the clinical note structure—different centers generate EHR databases according to different algorithms from different EHR software. UCSF colonoscopy reports begin with patient- and event-specific information, followed by medications, descriptions of procedure, colon findings, impressions and recommendations. Naturally, SFGH report contents are displaced differently as well.

Therefore, we presented ChatGPT with two versions of each report for our prompts: the first being the original, intact note, and the second, a note where the head of the report is trimmed at a unique string match of ";Procedure:" and trimmed at the tail at a unique string match of ";CPT". Similarly, for the SFGH set of reports, following a fixed structure for colonoscopy reports, we trimmed these notes at string matches of "INDICATIONS:" and "____". These report modifications led to a 14.3% and 15.9% average reduction in report characters for UCSF and SFGH, respectively.

***Prompt Development***

In our development phase we sampled 10 random notes each from both centers for validation, but report never having perfect classification accuracies in our prompt. We include colonoscopy reports that are not UC related since real-world colonoscopy reports from the EHR are not UC related. For example, our colonoscopy reports contain instances of patients with clear diagnoses of Crohn's disease (CD) or colon cancer. The motivation for this real-world sampling model of colonoscopy reports is to simulate the deployment of our protocol on an EHR database. A simple and fluid pipeline involves querying note text, extracting information with an LLM, and mapping extracted information uniquely to a patient-encounter identifier, effectively providing a measure of patient health at a given time point. We utilized UCSF's Versa application programming interface (API), UCSF's PHI compliant access to GPT-4, and sent prompts with the LangChain framework. Broadly used for high-power, client-facing applications, LangChain allows for context rich conversations and prompts in a programmatic way. As opposed to a very limited interactive chat interface, LangChain allows us to send hundreds of clinical reports within a single prompt context. In particular, our prompts followed the conversational templates in **Table 1** where "system" refers to setting a system tone for the AI; "human" refers to the prompts sent by the prompter; and "ai" refers to the responses produced by ChatGPT. Templating allows us to produce consistent, standardized responses from GPT-4. The prompts that we developed can also be found in **Table 1**. Little modification was performed from the original Silverman et al. annotation protocol (3). Words and phrases were added for clarity and text was modified for precision and occasionally for conciseness.

***Classification Assessment and Cost Calculation***

We measure the performance of each algorithm according to practical measures and standard statistical learning measures such as balanced accuracy, macro-averaged precision, recall and F1-score (1, 2). In particular, for practical measures we computed the MES scorable accuracy, determining if the report is

MES scorable; severity agreement, a measurement converting the ordinal MES {0,1,2,3} scale to {0,1} for *not* moderate-to-severe and moderate-to-severe, respectively; and under-classification(k) error, for the percentage of reports that were labeled k-points lower than the true label. These metrics were computed with scikit-learn (v1.4).

The average cost was computed by first counting token counts for sending prompts to GPT-4 32k token deployment. At the time of writing, sending prompts costs $0.06 per 1000 tokens, and receiving text responses, *completions*, costs $0.12 per 1000 tokens. We utilized the tiktoken (v0.6.0) library to compute token counts and costs. We show average token counts and 1.96 standard deviation spread for each prompt variation studied in **Table 2.**

***Software***

All scripts and calculations were run in Python 3.10.9.

**Table 1**. Zero-shot and N-shot prompt templates for interacting with ChatGPT through LangChain are presented. Variables such as “protocol”, “ explanation” and “colonoscopy_report” are filled according to our prompt parameters. The “ai” responses are produced from iteratively building on prompts preceding those responses.  Zero-shot, one-shot and four-shot protocols texts are shared under the same N-shot protocol template. We also show the binary Mayo scorable protocol as well as the strict Mayo scorable-only protocol.

| Prompt/ Protocol Type | Template/Text |
| --- | --- |
| Zero-Shot **Prompt Template** | [<br>("system", "You are a very competent, and currently board certified gastroenterologist. \\<br>You are incredibly adept with being able to understand colonoscopy procedure reports\\<br>from any medical center."),<br>("human", protocol)<br>] |
| N-Shot **Prompt Template** | [<br>("system", "You are a very competent, and currently board certified gastroenterologist. \\<br>You are incredibly adept with being able to understand colonoscopy procedure reports\\<br>from any medical center."),<br>("human", protocol),<br>("ai", "Thank you for providing these examples. I have reviewed them to understand the scoring system and\\<br>the specifics of annotation. Now I am ready to review and score any colonoscopy reports you provide\\<br>following the protocol you described."),<br>("human", "Of course, thank you for your help. I really appreciate it. Before I ask you to review any formal\\<br>reports, I'd like to first provide some example reports. Following are some example reports annotated by\\<br>humans according to the protocol I just described. I would like you to familiarize yourself with them. \\<br>In particular I am providing {sampset} notes from {centerLearn}, {samp} for each label of -1,0,1,2,3\\<br>according to the annotation protocol. You will see that for each '[Input k]' there is an associated\\<br>'[Output k]' corresponds to the correct label where 'k' corresponds to the example report 'k'. Here are the\\<br>notes: {report_sample}"),<br>("ai", "You've shared a significant amount of data, and I have analyzed it thoroughly. I'm now ready to examine any\\<br>colonoscopy reports you'd like me to review. Please provide the reports and I will apply the annotation\\<br>protocol to determine whether they can be annotated, and if so, assign them a Mayo score."),<br>("human", "Excellent. Thank you for telling me this. Now, with the entire protocol captured above, please\\<br>label/score the following colonoscopy report from {centerLabel}---please provide the label\\<br>with {explanation}: {colonoscopy_report}")<br>] |
| N-Shot (Five Class) **Protocol Text** | "Hello, I will provide you with some colonoscopy reports of patients. I would please like you to first determine\\<br>if a report can be annotated based on the following protocol:\\<br>- If a patient had a proctocolectomy the report should NOT be annotated (a total abdominal colectomy is\\<br>acceptable for annotation to enable scoring of the Hartmann's pouch).\\<br>- If a procedure had no mucosa visualized due to a poor bowel preparation the report should NOT be annotated.\\<br>- If a patient has a clear diagnosis of Crohn's disease the report should NOT be annotated.\\<br>- For example, Crohn's appears in the impressions section, or it appears in the indications section only and\\<br>the findings to do not seem to question diagnosis.\\<br>- If the patient carries a diagnosis of UC but has Crohn's disease like features (stricture, fistulae, aphthae), \\<br>and impressions section does not clearly indicate a revised diagnosis from ulcerative colitis \\<br>to Crohn's Disease, then the report CAN be annotated.\\<br>- If the patient carries a diagnosis of 'IBD' then the report CAN be annotated so long as there is no\\<br>mention of the patient having 'stricture(s)' and/or 'fistula(s)'. \\<br>In particular, I would like you label reports that cannot be annotated with a '-1'.\\<br>\\<br>If a report can be annotated, then you will score the report with a Mayo score. In particular, a Mayo score for\\<br>a patient with ulcerative colitis will be given a Mayo score according to the most severely affected segment of the\\<br>visualized colorectum according to the following descriptors:\\<br>- Please score a colonoscopy report with '0' for mentions of 'normal' (normal appearance),\\ |

| | |
|---|---|
| | 'quiescent', 'scar' without other descriptors in the colorectum consistent with other classes.\<br>- Please score a colonoscopy report with '1' for mentions of 'erythema', 'decreased vascular pattern',\<br>'granularity', 'aphthous ulcer', 'aphthae', 'mild', no friability \<br>- Please score a colonoscopy report with '2' for mentions of 'friability', marked or extensive 'erythema',\<br>'loss of vascularity (absence)', 'erosions' \<br>(these can adjective mentions additionally distinguished with 'moderate').\<br>- Please score a colonoscopy report with '3' for mentions of spontaneous 'bleeding', \<br>'ulcers/ulcerated/ulceration', or 'severe' conditions.\<br>\<br>Please be mindful of the following edge cases: \<br>- In the case of poor bowel prep only please score the report amongst the segments that are seen according\<br>to the previously mentioned scoring protocol.\<br>- If the the colonoscopy report indicates 'moderate-to-severe' but without any other descriptors of \<br>severity please score the report with '3'.\<br>- If a clinician assignment of a Mayo score is discordant with other descriptors (e.g., report describes\<br>severity as mild but indicates presence of an ulcer), please score the report with the most severe\<br>descriptor (e.g., 'severe colitis' with 'erosions' will be scored as '3', and 'mild' colitis with\<br>an 'ulcer' will be scored as '3'). \<br>- For mentions of incomplete colonoscopy, sigmoidoscopy or proctoscopy (e.g., Hartmann's pouch) if the exam\<br>was intentionally halted due to distal disease severity, please score as a 3. Otherwise, please score\<br>only the segments that are seen.\<br>- IBD patients with...\<br>- 'aphthous ulcers', please score with a '1'.\<br>- 'SCAD (erythematous mucosa), please score with a '1'\<br>- 'granular mucosa', please score with a '3'.\<br>- 'deep', 'shallow' or fissuring' ulcer,' please score with a '3'.\<br>- 'deep erosion', please score with a '2'.\<br>- 'scar/scarring', if described as associated with loss of vascularity, score with at least a '1'.\<br>Otherwise, the report cannot be annotated and should be labeled, '-1'.\<br>- 'mucosal healing', ignore this descriptor and look for other descriptors to classify the severity.\<br>- 'no active inflammation', ignore this descriptor and look for other descriptors to\<br>classify the severity.\<br>- If a patient does not a have a clear diagnosis of IBD (e.g., could be a screening colonoscopy or a\<br>diagnostic exam), so long as no exclusion criteria are met we can assign a Mayo score per the general\<br>rules (e.g., a clinical diagnosis of ulcerative colitis may be\<br>subsequently assigned)" |
| Mayo Scorable (Binary) **Protocol Text** | "Hello, I will provide you with some colonoscopy reports for patients. I would please like you to first\<br>determine if a report can be annotated based on the following protocol:\<br>- If a patient had a proctocolectomy the report should NOT be annotated (a total abdominal colectomy is\<br>acceptable for annotation to enable scoring of the Hartmann's pouch).\<br>- If a procedure had no mucosa visualized due to a poor bowel preparation the report should NOT be annotated.\<br>- If a patient has a clear diagnosis of Crohn's disease the report should NOT be annotated.\<br>- For example, Crohn's appears in the impressions section, or it appears in the indications section only and\<br>the findings to do not seem to question diagnosis.\<br>- If the patient carries a diagnosis of UC but has Crohn's disease like features\<br>(stricture, fistulae, aphthae), and impressions section does not clearly indicate a revised diagnosis from \<br>ulcerative colitis to Crohn's Disease, then the report CAN be annotated.\<br>- If the patient carries a diagnosis of 'IBD' then the report CAN be annotated so long as there is no mention\<br>of the patient having 'stricture(s)' and/or 'fistula(s)'. \<br>- IBD patients with mentions of 'scar/scarring', if described as associated with loss of vascularity, \<br>score with '1'. Otherwise, the report cannot be annotated and should be labeled, '0'. \<br>\<br>In particular, I would like you label reports that cannot be annotated with a '0' and reports that can be\<br>annotated with '1'. Please label the following note (please provide {explanation}): {colonoscopy_report}" |
| Mayo Scoring (4 Class) **Protocol Text** | "Hello, I will provide you with some colonoscopy reports for patients. The notes are Mayo scorable. In \<br>particular, a Mayo score for a patient with ulcerative colitis will be given a Mayo score \<br>to the most severely affected segment of the visualized colorectrum according to the following descriptors:\<br>- Please score a colonoscopy report with '0' for mentions of 'normal' (normal appearance), 'quiescent',\<br>'scar' without other descriptors in the colorectum consistent with other classes.\<br>- Please score a colonoscopy report with '1' for mentions of 'erythema', 'decreased vascular pattern',\<br>'granularity', 'aphthous ulcer', 'aphthae', 'mild', no friability \<br>- Please score a colonoscopy report with '2' for mentions of 'friability', marked or extensive 'erythema',\<br>'loss of vascularity (absence)', 'erosions'\<br>(these can adjective mentions additionally distinguished with 'moderate').\<br>- Please score a colonoscopy report with '3' for mentions of spontaneous 'bleeding', \<br>'ulcers/ulcerated/ulceration', or 'severe' conditions.\<br>\ |

| | |
|---|---|
| | Please be mindful of the following edge cases: \ <br>- In the case of poor bowel prep only please score the report amongst the segments that are seen according to\ <br>the previously mentioned scoring protocol.\ <br>- If the the colonoscopy report indicates 'moderate-to-severe' but without any other descriptors of severity\ <br>please score the report with '3'.\ <br>- If a clinician assignment of a Mayo score is discordant with other descriptors (e.g., report describes\ <br>severity as mild but indicates presence of an ulcer), please score the report with the most severe\ <br>descriptor (e.g., 'severe colitis' with 'erosions' will be scored as '3', and 'mild' colitis with an\ <br>'ulcer' will be scored as '3'). \ <br>- For mentions of incomplete colonoscopy, sigmoidoscopy or proctoscopy (e.g., Hartmann's pouch) if the exam was\ <br>intentionally halted due to distal disease severity, please score as a 3. Otherwise, please score only\ <br>the segments that are seen.\ <br>- IBD patients with...\ <br>- 'aphthous ulcers', please score with a '1'.\ <br>- 'SCAD (erythematous mucosa), please score with a '1'\ <br>- 'granular mucosa', please score with a '3'.\ <br>- 'deep', 'shallow' or fissuring' ulcer,' please score with a '3'.\ <br>- 'deep erosion', please score with a '2'.\ <br>- 'scar/scarring', if described as associated with loss of vascularity, score with at least a '1'.\ <br>- 'mucosal healing', ignore this descriptor and look for other descriptors to classify the severity.\ <br>- 'no active inflammation', ignore this descriptor and look for other descriptors to classify the severity.\ <br>- If a patient does not a have a clear diagnosis of IBD (e.g., could be a screening colonoscopy or a diagnostic\ <br>exam), so long as no exclusion criteria are met we can assign a Mayo score per the general rules\ <br>(e.g., a clinical diagnosis of ulcerative colitis may be subsequently assigned)\ <br>\ <br>With the entire protocol captured above, please label/score the following colonoscopy report---please provide the\ <br>label with {explanation}: {colonoscopy_report}" |

**Table 2**. Results of each prompt and costs for each API call according to each prompt variation.

| Prompt Template Variation | Zero-shot | Zero-shot | Zero-shot | Zero-shot | Zero-shot | Zero-shot | Zero-shot | Zero-shot | Zero-shot-1, Binary Class | Zero-shot-1, Binary Class | Zero-shot-2, Mayo Score | Zero-shot-2, Mayo Score | One-shot | One-shot | Four-shot | Four-shot |
|---|---|---|---|---|---|---|---|---|---|---|---|---|---|---|---|---|
| Center | UCSF | UCSF | SFGH | SFGH | UCSF | UCSF | SFGH | SFGH | UCSF | SFGH | UCSF | SFGH | UCSF | SFGH | UCSF | SFGH |
| With Parsing | Trimmed | Trimmed | Trimmed | Trimmed | Original | Original | Original | Original | Trimmed | Trimmed | Trimmed | Trimmed | Trimmed | Trimmed | Trimmed | Trimmed |
| With Explanation | Yes | No | Yes | No | Yes | No | Yes | No | Yes | Yes | Yes | Yes | Yes | Yes | Yes | Yes |
| Prompt Token Count | 1621.15 ±216.35 | 1606.15 ±216.35 | 2159.71 ±489.78 | 2144.71 ±489.78 | 1806.34 ±232.58 | 1794.34 ±232.58 | 2545.25 ±538.70 | 2530.25 ±538.70 | 951.15 ±110.38 | 1489.71 ±249.89 | 1340.16 ±110.38 | 1878.71 ±249.89 | 4574.16 ±110.39 | 8184.71 ±249.89 | 13339.16 ±110.38 | 24709.71 ±249.89 |
| Completion Token Count | 40.83 ±21.73 | 1.0 | 49.43 ±26.62 | 1.0 | 42.77 ±21.68 | 1.0 | 47.56 ±26.24 | 1.0 | 54.24 ±28.01 | 62.40 ±26.99 | 50.63 ±22.94 | 49.55 ±20.76 | 53.83 ±23.91 | 57.46 ±26.09 | 58.07 ±23.06 | 53.88 ±21.46 |
| Average cost per note | $0.097 | $0.096 | $0.129 | $0.128 | $0.108 | $0.107 | $0.152 | $0.1511 | $0.063 | $0.096 | $0.086 | $0.118 | $0.499 | $0.280 | $0.803 | $1.485 |
